# The impact of remote home monitoring of people with COVID-19 using pulse oximetry: a national population and observational study

**DOI:** 10.1101/2021.12.10.21267613

**Authors:** Chris Sherlaw-Johnson, Theo Georghiou, Steve Morris, Nadia E Crellin, Ian Litchfield, Efthalia Massou, Manbinder S Sidhu, Sonila M Tomini, Cecilia Vindrola-Padros, Holly Walton, Naomi Fulop

## Abstract

**Background:** Remote home monitoring of people testing positive for COVID-19 using pulse oximetry was implemented across England during the Winter of 2020/21 to identify falling blood oxygen saturation levels at an early stage. This was hypothesised to enable earlier hospital admission, reduce the need for intensive care and improve survival. This study is an evaluation of the clinical effectiveness of the pre-hospital monitoring programme, COVID oximetry @home (CO@h).

**Methods:** We analysed relationships at a geographical area level between the extent to which people aged 65 or over were enrolled onto the programme and outcomes over the period between November 2020 to February 2021

**Findings:** For every 10% increase in coverage of the programme, mortality was reduced by 2% *(95% confidence interval: -4% to 1%)*, admissions increased by 3% *(-1% to 7%)*, in-hospital mortality fell by 3% *(-8% to 3%)* and lengths of stay increased by 1·8% *(-1·2% to 4·9%)*. None of these results are statistically significant.

**Interpretation:** There are several possible explanations for our findings. One is that the CO@h did not have the hypothesised impact. Another is that the low rates of enrolment and incomplete data in many areas reduced the chances of detecting any impact that may have existed. Also, CO@h has been implemented in many different ways across the country and these may have had varying levels of effect.

**Funding:** This is independent research funded by the National Institute for Health Research, Health Services & Delivery Research programme (RSET Project no. 16/138/17; BRACE Project no. 16/138/31) and NHSEI. NJF is an NIHR Senior Investigator. The views expressed in this publication are those of the authors and not necessarily those of the National Institute for Health Research or the Department of Health and Social Care.

**Research in Context:** *Evidence before this study:* Existing evidence before this study and the search strategy used to obtain this evidence has been published previously by the authors in a systematic review. Previous quantitative studies have assessed remote oximetry monitoring services for COVID-19 patients mostly at individual sites and focussed on their safety. However, their effectiveness has been little studied. This may reflect the challenges of identifying reliable counterfactuals during a rapidly evolving pandemic.

*Added value of this study:* This study is part of a wider mixed methods evaluation that followed the rapid implementation of remote monitoring across the English NHS during the Winter of 2020/21. It adds to the evidence of the effectiveness of such programmes at a national level.

*Implications of the available evidence:* There is some existing evidence that remote monitoring of COVID-19 patients can be locally effective although we have not been able to replicate such findings at a wider level. Missing data and lower coverage of the service than expected may have influenced our results, and the effectiveness of some local programmes could have been lost among the analysis of national data. Future implementation requires better data collection strategies which could be focussed within fewer local areas, and effective learning from areas that have achieved better population coverage.

## Background

During the early months of the COVID-19 pandemic, many patients with COVID-19 were admitted to hospital having deteriorated several days after they were first diagnosed. Many of these patients had “silent hypoxia” (low blood oxygen saturation levels without typical symptoms or awareness) and, once at hospital, often required intensive treatment with a high risk of mortality.^1^ This motivated health services to try and detect such cases at an earlier stage by monitoring blood oxygen levels in people diagnosed with COVID-19 at home using pulse oximetry. This could reassure people who did not need to go to hospital, whilst more quickly identifying individuals with dangerously low blood oxygen saturations (<92%).^2,3^

In the English National Health Service (NHS), remote home monitoring using pulse oximetry started to be implemented within some areas during the first wave of the pandemic in the UK.^4^ This was followed by a national implementation during the Winter of 2020/21.^5^ The service was known as COVID Oximetry @home (CO@h) and by the end of January 2021 it was operating in all clinical commissioning areas of England.

The way different areas organised and operated the service varied. People testing positive for COVID-19 would be sent a pulse oximeter for use at home and readings would be sent to local healthcare staff. The process of reporting readings was sometimes facilitated by smartphone technology or reported via telephone, depending on the location and the preferences of the patient.^4^ Some sites started by only enrolling individuals aged 65 or over, or who were deemed extremely clinically vulnerable. Others extended enrolment to a wider age group, and often these criteria changed over time.^6^

One aim of CO@h was to reduce mortality through earlier identification of deterioration. Furthermore, it was hypothesised that if fewer COVID-19 patients were admitted to hospital with advanced disease, and critically low oxygen levels, there may be a reduction in the use of critical care facilities, fewer deaths within hospital and shorter lengths of stay. The anticipated impact on numbers of hospital admissions was less certain since the aim of the programme was not to reduce admissions, but to make sure people who needed to be in hospital were admitted sooner. However, any consequence on the number and mix of patients admitted for COVID-19 would be useful to understand as remote monitoring may have different impacts on different types of individual.

Earlier studies of the use of oximetry for remote monitoring within England during the country’s first wave focussed on aspects of safety and implementation, but were unable to establish reliable comparators for measuring impact.^7,8^

Faced with this lack of evidence as to the likely effectiveness of CO@h, the two rapid evaluation teams commissioned by the National Institute for Health Research (NIHR) were requested by NHS England to undertake a mixed methods study of the service.^9^ This study included evaluations of clinical effectiveness, costs, the processes of implementation and patient and staff experiences, and was one of three evaluations simultaneously requested by NHS England.^9–11^

This paper presents findings from the clinical effectiveness workstream of the study addressing the specific research questions:

1. What is the impact of CO@h on mortality?
2. What is the impact of CO@h on the incidence of hospital admission for COVID-19 or suspected COVID-19 and on the characteristics of those admitted?
3. For these admissions, what is the impact on in-hospital mortality and length of stay?

Our quantitative approach used combinations of unlinked, aggregated population-level data and hospital administrative data. In doing so we were able to undertake a rapid analysis that not only complemented the other evaluations but provided valuable insight in the future evaluation of similar programmes implemented at scale.

## Methods

### Study design

The study of overall mortality and admissions was designed as an area-level analysis combining aggregated data from different sources. Considering these data as time series, we investigated “dose-response” relationships^12^ between the evolving coverage of the programme within each area and outcome. We analysed four outcomes: mortality from COVID-19, hospital admissions for people with confirmed or suspected COVID-19, in-hospital mortality for these admissions and their lengths of stay. For the in-hospital outcomes, we used an observational design relating in-hospital mortality and lengths of stay at an individual patient level to the degree of coverage of the CO@h programme within the area at the time of admission.

### Setting and participants

The setting was all Clinical Commissioning Group (CCG) areas in England where there was complete data on the number of people enrolled onto the programme (onboarded) between 2^nd^ November 2020 and 21^st^ February 2021. (CCGs are NHS organisations that organise the delivery of primary care services within a specific geographic area. At the time of the study there were 135 in England.) The study populations included anyone with a laboratory-confirmed positive test for COVID-19 and any hospital admission for COVID-19 or suspected COVID-19. We also limited the analysis to people aged 65 or over, as this population was eligible for CO@h across all CCGs and both coverage and frequency of outcomes within this group were higher. Implementation among younger age groups across the country was much more variable.

### Data and variables

For our analysis we used data from several sources (see supplementary material). Data on numbers of new cases of COVID-19 and deaths were acquired from Public Health England (now the UK Health Security Agency). New cases were laboratory-confirmed and deaths were those either within 60 days of the first positive test or where COVID-19 was mentioned on the death certificate.^13^ If someone had more than one positive test within the previous seven days, then only one was counted.^14^ These data were aggregated by week, age band and CCG. The selected age bands were 65 to 79 and 80 plus. Numbers of people onboarded to CO@h were sourced from a bespoke national data collection for the programme and aggregated by the team at Imperial College London undertaking one of the other two simultaneous evaluations. Due to small numbers, aggregation was performed by fortnight, rather than week, and by the same age bands and by CCG. To comply with data protection rules, these data were also rounded to the nearest five individuals, or, for smaller values, labelled as between one and seven.

Data on hospital admissions and outcomes were obtained from Hospital Episode Statistics (HES). Although most of the non-hospital data was available weekly, we aggregated to fortnightly data in order to match the aggregation of the onboarding data. We restricted our statistical analysis to the period between 2 November 2020 and 21 February 2021 when numbers of cases and outcomes were at their peak. Also, outside that period there were too many low numbers at our chosen level of granularity.

Coverage of CO@h was measured as numbers enrolled onto the programme within each CCG every fortnight divided by the number of new cases detected in that fortnight. To be able to calculate this by CCG, we required the onboarding data within a CCG to be complete. CCGs providing complete onboarding data were identified by NHS Digital. As part of the wider mixed methods study, the team selected 28 study sites for surveys, interviews and to obtain data on costs, most of which were CCGs that provided complete data. For the costing part, sites were independently asked how many people they had onboarded, and we used this information to validate the reports of completeness from the national programme and to include additional CCGs where the numbers onboarded were broadly similar or greater. Further information about this process is included in the supplementary material. Where numbers onboarded were between one and seven, we assigned a value of four, being the mid-point within the range.

We estimated coverage in two ways. One was to calculate it for each CCG regardless of whether a service was operating at the time, and this was used in our analysis. However, to understand what coverage was achievable once a service was implemented, we also estimated coverage within individual CCGs over periods when we knew a service was operating there. For this we only included fortnights over which a service was operating within the CCG for the entirety.

The proportion of hospital beds occupied by COVID-19 patients was used as a measure of local system pressures and sourced from publicly available routine data.^15^ By the end of February 2021, most hospital trusts were operating step-down virtual wards whereby COVID-19 patients could be discharged early with a pulse oximeter and monitored at home in a similar way to the CO@h service.^16^ Due to the potential influence of these virtual wards on hospital outcomes their existence was incorporated as a confounding variable in our analyses of length of stay and in-hospital mortality.

### Comparisons between included and excluded CCGs

We compared population characteristics and COVID-19 incidence rates between the CCGs we included, because their data was believed to be complete, and the remaining CCGs to test how representative the included CCGs were. The mean values and proportions associated with each CCG were treated as the separate observations and comparisons were carried out using Student t-test, or Mann-Whitney U-tests where data were skewed. We also investigated their geographical spread.

### Analysis of mortality

Because we only had aggregate data for deaths, new COVID cases and people onboarded to CO@H, our approach was to calculate coverage rates for CO@H over time and then investigate relationships between levels of coverage and mortality by age band within each CCG. To do this we adopted a two-stage approach. The first stage was to estimate denominators representing exposure, the second was to use these as offset variables in negative binomial regression models, relating mortality to coverage of the CO@H programme by age group. We included a further variable for the month to allow for changes in relationships as the second wave progressed. To account for CCG-level effects we used general estimating equation (GEE) approaches.^17^ This approach accommodates the fact that mortality within a single CCG is likely to be correlated and GEEs ensure that correlation is accounted for by adjusting parameter estimates and standard errors.

The need to estimate denominators arose because we were not able to directly link the new cases and mortality data. When a death occurs, the median time between a new case arising and death is about two weeks, although some may have been diagnosed only in the previous week, and some three weeks or more before. We therefore developed a preliminary set of regression models relating mortality to new cases, with new cases lagged at different times, in order to establish the contributions of the lagged variables. These then determined weights which we used to aggregate new cases into a denominator. Assuming that there was no lag between diagnosis and exposure to the programme, we applied the same weights to the onboarding data to establish a weighted coverage variable appropriate to the mortality observed at each time. A more detailed description of this approach is provided in the supplementary material.

Other options for lagging the time between diagnosis, onboarding and mortality were tested in sensitivity analysis and reported in the supplementary material.

### Analysis of hospital admissions

Hospital admissions over the study period were extracted from Hospital Episode Statistics (HES). We considered any admission where COVID-19 or suspected COVID-19 appeared as a diagnosis in the first episode of care, whether as a primary or secondary diagnosis (ICD-10 codes U07.1 and U07.2). If a patient was readmitted with one of these diagnoses within a 28-day period, we only considered the first admission. To match the onboarding data, numbers were aggregated by age band and fortnight.

We undertook a similar procedure for hospital admissions as for mortality, although with different weights, since the time between diagnosis and admission tended to be shorter.

Again, for our sensitivity analysis, we tested different options for lagging the time between diagnosis, onboarding and outcomes. We also tested the option of only including admissions where COVID-19 or suspected COVID-19 was the primary diagnosis.

Separate models were developed to evaluate any impact of CO@h on the characteristics of patients admitted in terms of age, sex, deprivation, Charlson Score (a measure of the severity of co-morbidities) and ethnicity, controlling for month and accounting for CCG-level effects as before.

### Analysis of In-hospital outcomes

To analyse outcomes for patients admitted to hospital, we used individual-level Hospital Episode Statistics (HES). To investigate the impact on in-hospital mortality, we created logistic regression models relating mortality to the weighted coverage for the relevant CCG with individual patient characteristics as confounders. Values for the weighted coverage corresponded to those calculated for hospital admissions. Again, we used general estimating equation (GEE) approaches to account for CCG-level effects. For length of stay we used negative binomial regression models^18^ with stays longer than 30 days trimmed to 30 days. For this analysis, length of stay was defined as the number of days between admission and discharge from the same hospital or death within that hospital.

### Using rounded data

To accommodate the uncertainty caused by the rounding of the onboarding data, we ran all our statistical models multiple times, each time randomly sampling onboarded numbers from the range of feasible values (treating the distributions as uniform). Based on the similarity of results with each simulation, we deemed it sufficient to perform 1000 runs for each model. The simulation results were then pooled to obtain overall effect sizes. All statistical analyses were performed using SAS version 9.4.^19^

### Patient and Public Involvement

Members of the study team met to discuss the study with service users and public members of the NIHR BRACE Health and Care Panel and patient representatives from NIHR RSET. Although mostly used for the qualitative evaluations in the wider study, meetings were held during the data analysis phase to share learning and cross-check our interpretations of findings.

### Data governance and ethics

The receipt of aggregated data from Public Health England was governed by a data sharing agreement. Receipt of aggregated onboarding data from Imperial College was governed by their separate data sharing agreement with NHS Digital. The access and use of HES was governed by an existing data sharing agreement with NHS Digital covering NIHR RSET analysis (DARS-NIC-194629-S4F9X). Since we were using combinations of aggregated data and datasets for which we already had approval to use, no ethics committee approval was needed for this analysis.

## Results

### Data completeness and coverage

Over the period of analysis, we judged that onboarding data was complete for 37 CCGs (27% of the total number of 135 CCGs across England).

The included CCGs had no notable differences in mean age, proportions of non-White population or proportions resident in most deprived areas when compared to the remaining 98 that were not included; although included CCGs had a lower incidence of positive test results (Table 1). There were also regional differences: no CCGs from the East NHS Region were included, and only one from the North East and Yorkshire (Figure 1). The South West, North West and Midlands were the best represented regions.

**Table 1.** Characteristics of the populations resident within the CCGs included in the analysis compared with those that were excluded. (Samples are the proportions and rates observed within each CCG).

|  | Included CCGs (n=37) |  | Excluded CCGs (n=98) |  | P-value for difference between groups* |
| --- | --- | --- | --- | --- | --- |
|  | Mean | (Standard error) | Mean | (Standard error) |  |
| Mean proportion aged 65 or more | 18.1% | (0.7%) | 17.9% | (0.5%) | 0.77 |
| Mean proportion aged 80 or more | 4.8% | (0.2%) | 4.8% | (0.1%) | 0.91 |
| Mean proportion in non-white ethnic groups | 17.3% | (2.7%) | 17.7% | (1.8%) | 0.97 |
| Mean CCG-level Index of Multiple Deprivation | 22.7 | (1.3) | 22.6 | (0.8) | 0.96 |
| Incidence of laboratory-confirmed COVID-19 per 1000 people |  |  |  |  |  |
| All ages | 41.9 | (1.7) | 46.8 | (1.4) | 0.03 |
| Age 65 or over | 30.4 | (1.3) | 35.0 | (1.2) | 0.01 |

\*Two-sample T-tests. Mann-Whitney test for ethnic groupings

**Figure 1.**
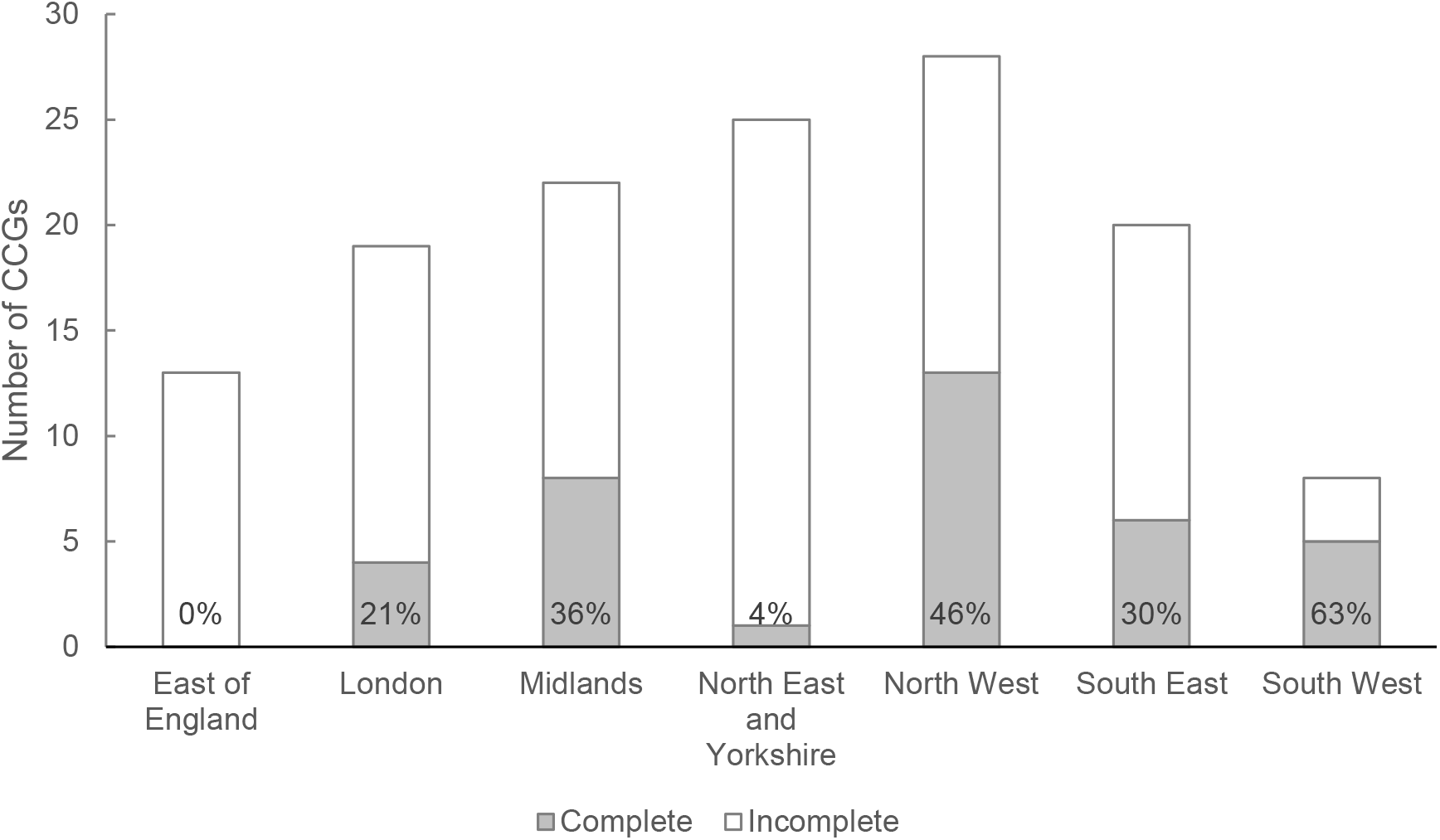
Number of CCGs with complete data by NHS region (% complete shown in each bar)

Fortnightly coverage of CO@h among people aged 65 or over within the 37 CCGs is shown in Figure 2. Rates were particularly low earlier in the period because many sites had not commenced implementation. The numbers of CCGs where there were sites onboarding patients within each fortnight is shown along the horizontal axis. Sites within nine (24%) CCGs were operating services in the first fortnight, which had risen to 33 (89%) within the fortnight beginning 28 December. Services were operating within all CCGs during the fortnight beginning 11 January 2021. The median coverage only exceeded 10% in the final fortnight, although, from the end of November, the maximum was consistently above 30%, with one or two CCGs each fortnight achieving much higher rates than the rest. The overall coverage over the period across all 37 CCGs was 5.9%. If we exclude fortnights during which services were either not operating or operating for only part of the fortnight, the overall coverage was 8.7% with only one CCG averaging a rate of more than 30%.

**Figure 2.**
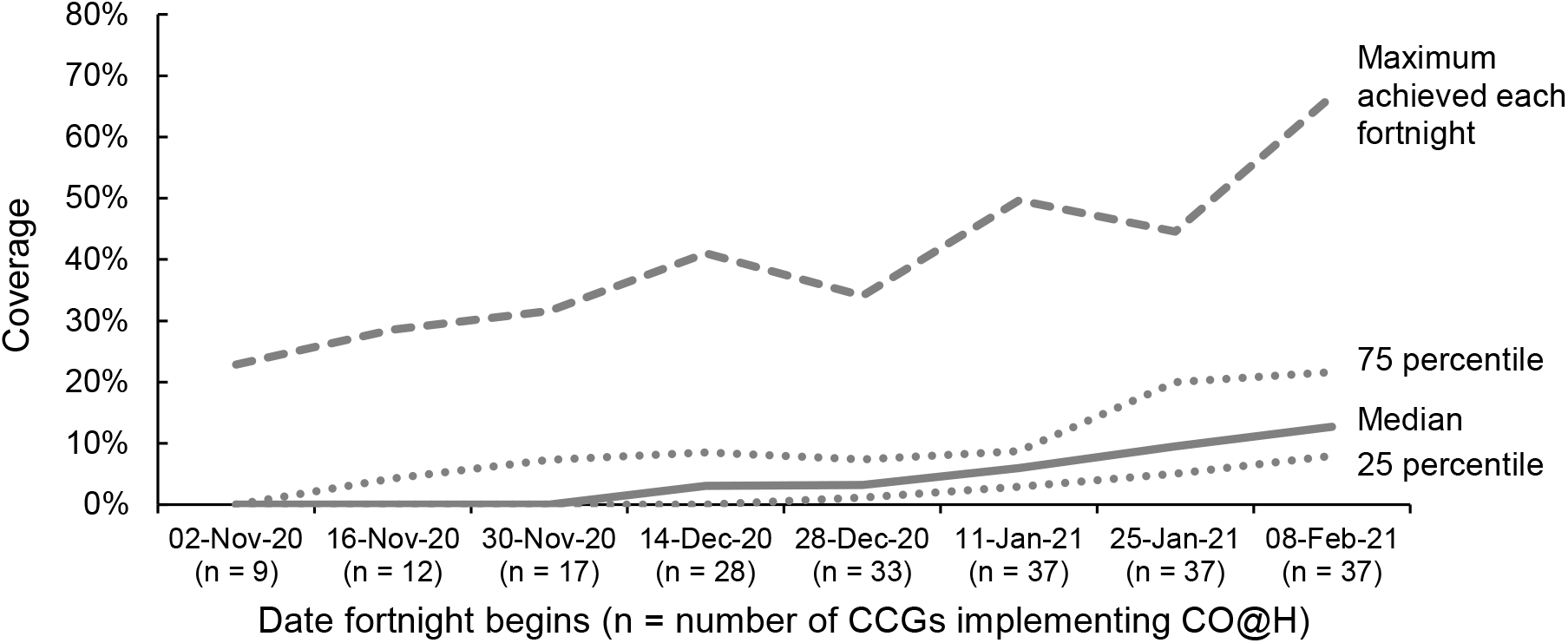
Variation in coverage of CO@h among people aged 65 or over with laboratory-confirmed COVID-19 across the 37 CCG’s included in the analysis.

### Summary outcomes

Summary outcomes for the period from 2 November 2020 to 21 February 2021 for the 37 CCGs by age band are shown in Table 2. Although there were many more positive tests recorded among the under 65’s, there were approximately half as many deaths and equivalent numbers of hospital admissions. Coverage after implementation was low across both age groups and highest among those aged 65 to 79.

**Table 2.** Outcomes across the 37 CCGs included in the analysis between 2nd November 2020 and 21 February 2021.

|  | Age band |  | Both age groups |
| --- | --- | --- | --- |
|  | 65 to 79 | 80+ |  |
| Positive tests and onboarding over the whole period |  |  |  |
| Number of new positive diagnoses | 41,437 | 27,944 | 69,381 |
| Number onboarded to CO@h* | 2,734 | 1,382 | 4,116 |
| Coverage | 6.60% | 4.95% | 5.93% |
| Positive tests and onboarding since implementation** |  |  |  |
| Number of new positive diagnoses | 26,507 | 18,636 | 45,143 |
| Number onboarded to CO@h* | 2,610 | 1,320 | 3,930 |
| Coverage | 9.85% | 7.08% | 8.71% |
| Mortality |  |  |  |
| Number of deaths | 4,269 | 8,699 | 12,968 |
| Hospital admissions |  |  |  |
| Number of hospital admissions | 12,351 | 12,128 | 24,479 |
| Number of hospital admissions (excluding readmissions within 28 days) | 10,895 | 10,683 | 21,578 |
| Within hospital outcomes |  |  |  |
| Number of in-hospital deaths | 2,748 | 3,948 | 6,696 |
| In-hospital deaths per admission | 22.25% | 32.55% | 27.35% |
| Median length of stay in days (*IQR*) | 8 (*3 to 15*) | 9 (*4 to 17*) | 8 (*4 to 16*) |
| Number staying 2 weeks or more | 3,489 | 4,134 | 7,623 |
| Proportion staying 2 weeks or more | 28.25% | 34.09% | 31.14% |

\*Onboarded numbers aggregated from fortnightly counts at CCG level rounded to the nearest 5 or given the value 4 if between 1 and 7.
\*\*Only includes fortnights for which a service was implemented within the CCG over the whole period

### Mortality and hospital admissions

Results from our models for mortality and hospital admission are shown in Table 3. For every 10% increase in coverage, mortality fell by 2% (relative risk = 0.98, 95% confidence interval: 0.96 to 1.01) and admissions increased by 3% (relative risk = 1.03, 95% confidence interval: 0.99 to 1.07), but neither result is statistically significant. There is, however, a significantly higher risk of mortality and admission among the older age group and higher risk of mortality in the months following November 2020.

**Table 3.**
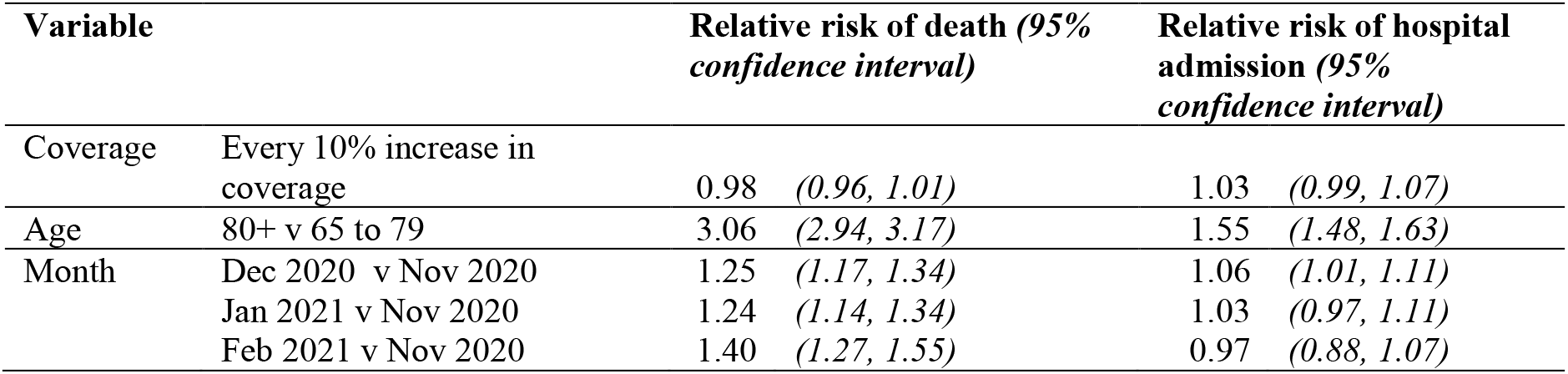
Results of the multivariate analysis for mortality and hospital admission: relative risks associated with each factor in the model

We also found no significant relationship between the coverage of CO@h and the ethnicity, sex, deprivation or health status of patients admitted to hospital with COVID-19 or suspected COVID-19 (Table 4). The impact on age is of borderline statistical significance with each 10% increase in coverage associated with a reduction of nearly 4 months (p = 0.07).

**Table 4.** Impact of increases in coverage of oximetry on the characteristics of people admitted to hospital with COVID-19 or suspected COVID-19

| Characteristic | % change with each 10% increase in coverage (95% confidence interval) |
| --- | --- |
| Proportion non-white ethnicity | 0.43% (-8.99%, 10.82%) |
| Proportion female | 0.07% (-1.93%, 2.11%) |
| Proportion resident in most deprived areas (by quintile) | 0.30% (-7.19%, 8.41%) |
| Proportion with Charlson scores > 5 | -2.75% (-6.37%, 1.00%) |
| Change in mean age with each 10% increase in coverage (95% confidence interval) |  |
| Mean age | -3.9 months (-7.9, 0.2) |

### In-hospital outcomes

The results of our analysis of in-hospital outcomes are shown in Table 5. For every 10% increase in coverage, in-hospital mortality fell by 3% (relative risk = 0.97, 95% confidence interval: 0.92 to 1.03) and length of stay increased by 1.8% (95% confidence interval: -1.2% to 4.9%). Again, neither result is statistically significant. Non-White ethnicity and existing COVID bed occupancy on admission were both associated with shorter lengths of stay.

**Table 5.** Results of the multivariate analysis for in-hospital mortality and length of stay. Effects of each factor on the odds of mortality and proportionate change in length of stay.

| Variable |  | Odds ratio associated with in-hospital mortality (95% confidence interval) | Relative change in length of stay (95% confidence interval) |
| --- | --- | --- | --- |
| Coverage | Every 10% increase in coverage | 0.97 (0.92, 1.03) | 1.8% (-1.2%, 4.9%) |
| Age | 80+ v 65 to 79 | 1.73 (1.63, 1.84) | 6.5% (3.1%, 9.7%) |
| Month | Dec 2020 v Nov 2020 | 1.27 (1.14, 1.42) | -4.2% (-9.5%, 1.5%) |
|  | Jan 2021 v Nov 2020 | 1.12 (1.00, 1.26) | -11.5% (-14.9%, -8.1%) |
|  | Feb 2021 v Nov 2020 | 0.88 (0.76, 1.03) | -19.7% (-27.4%, -11.1%) |
| Sex | Female | 0.71 (0.66, 0.76) | 2.5% (0.0%, 5.0%) |
| Charlson score | Greater than 5 | 2.14 (1.96, 2.33) | 1.1% (-2.2%, 4.6%) |
| Ethnicity | Non-white ethnicity | 1.24 (1.07, 1.42) | -12.0% (-16.2%, -7.6%) |
| Deprivation | Lowest IMD quintile | 0.98 (0.89, 1.07) | 0.0% (-3.0%, 3.1%) |
| Has a virtual ward |  | 0.96 (0.88, 1.04) | 1.0% (-6.5%, 9.1%) |
| COVID bed occupancy in the trust on admission | Every 10% increase in occupancy | 1.04 (0.97, 1.11) | -5.2% (-7.6%, -2.7%) |

### Sensitivity analysis

Results from our sensitivity analyses are shown in the supplementary material. None of the changes we made in our assumptions affected our findings with respect to the association between coverage of remote oximetry and outcomes.

## Discussion

### Summary of findings

In this study we have found no association between the coverage of pulse oximetry, as implemented by the CO@h programme, and COVID-associated mortality or COVID-associated admission to hospital. For such hospital admissions, we found no relationship to in-hospital mortality or length of stay. Also, for COVID-associated admissions, there appears to have been no associated change in the patient characteristics we have tested.

### Interpretation

There are several possible explanations for these results, and it would be premature to conclude that the COVID oximetry @home programme has had no impact. Firstly, limitations in data completeness meant that we were only able to analyse onboarding data for one quarter of CCGs across England. This, combined with lower than expected coverage within these CCGs, meant that our ability to detect any possible impact was smaller than anticipated. This also meant that some areas of the country were under-represented. Secondly, individual level associations may not have been seen at the aggregate level. Thirdly, qualitative findings from our wider study revealed that COVID oximetry @home was implemented in a variety of ways by different services within CCGs, some of which may have had more impact on outcomes than others.^6^ The possible impact of the intervention on hospital admissions was always uncertain, since the aim of the programme was to ensure people who needed hospital treatment were admitted at the right time. Also, once in hospital, the determinants of length of stay are complex and multifactorial, and may have varied during the time course of the second wave of COVID-19 infection in England.

### Strengths and limitations

The anticipated value of our approach with aggregated data was that it would complement the other two simultaneous quantitative evaluations of CO@h,^10,11^ could be carried out more rapidly and, if COVID-19 continues to stretch national health services, it could be more readily repeated as new data become available, provided the right information is routinely collected at source.

We were able to handle small number suppression and the rounding of aggregate data by multiple random sampling throughout the range of possible values and it was encouraging to discover that this uncertainly did not have a large impact on results.

Using aggregated data has some limitations because it does not enable us to trace direct links between the onboarding of an individual and their outcomes. There may also be an ecological fallacy, where individual-level effects are not observable in the aggregated data.^20^ However, obtaining linked individual-level data is a complex and potentially long process that may not always be feasible when there is a need to provide rapid feedback to a developing programme and where resources are stretched. Unfortunately, however, our ability to provide rapid feedback was compromised by delays in obtaining onboarding data which proved an understandable challenge for local services in the midst of a pandemic.

This has been part of a larger mixed methods study that has added insight into some of our findings and provided locally collected bespoke data against we could verify information we received centrally about coverage and data completeness. However, these checks could only be made against the 28 study sites and we were not able to verify the data from the other CCGs in the same way. Findings from surveys and interviews have helped interpret what we have found and, conversely, our data analyses have helped provide a context against which to understand the relative importance of the qualitative findings.^6^

We anticipated that finding a suitable comparator group during the national implementation of a programme was likely to be problematic, and we therefore avoided this problem by treating the relationship between coverage and outcome as a dose-response. However, the power to detect any impact in such an analysis depends on the level of coverage which, in practice, was lower than we hoped.

During the period of our analysis the vaccination programme was starting, and by the end of our study period 88% of people aged 65 or over had had at least one dose.^21^ Although this study investigates outcomes of people after being diagnosed with COVID-19, there is evidence that vaccination changes the subsequent risks of mortality and hospital admission^22^ which could have had a confounding effect on our analysis.

In the context of fortnightly data, we assumed a minimal lag between the diagnosis of COVID-19 and onboarding to CO@h. However, there was evidence from sites that they sometimes encountered delays in identifying positive cases,^6^ although the overall impact on this assumption is uncertain.

### Comparison with other studies

Prior to this study, very little was known about the quantitative impact of the use of pulse oximeters for remote home monitoring of people diagnosed with COVID-19. One of the other evaluations of the CO@h programme in England (not yet peer-reviewed) also found no significant impact on mortality or health service utilisation.^10^ However, the study did find reductions in mortality and increases in hospital attendance (yet with lower use of critical care) among people enrolled onto the programme after attending the Emergency Department (ED).^23^ A study of 4,384 high risk patients receiving home monitoring of vital signs, including pulse oximetry, in one region of Galicia, Spain, found lower admissions, lengths of hospital stays and in-hospital mortality when compared with other local regions.^24^ A recent study of CO@h carried out at one site demonstrated reductions in 30-day mortality and lengths of stay among people admitted to hospital.^25^ This, however, is currently a pre-print prior to peer review and lacks some details about the comparability of the control group. In another study implemented in the UK during the first wave, patients with suspected COVID-19 attending ED were discharged home with an oximeter. They observed a reattendance rate of 4.7% compared to 22.7% among a retrospective control group.^26^ However, this was a younger cohort (median age 41 years) and the absolute numbers of reattendance were small (nine in all). Other studies have reported on the safety of similar programmes, but have lacked comparators.^8,27–30^

### Implications and further research

At the start of this study we anticipated the services would have higher coverage and complete data would be available from more CCGs. Although the use of aggregated population-level data can enable more rapid evaluation of a new service, these two elements had an influence on the power of the analysis to detect an impact. The resulting shortfall in expected data reflects the challenges of trying to centrally manage a bespoke data collection while services are already stretched. However, sufficient quantities of data are vital to determining whether a service is effective, so it is important to understand how this can be improved, for example, by concentrating data collection in a few sites and using routinely collected data wherever possible.

Furthermore, low coverage raises questions about capacity of both staff and resources in the midst of high infection rates and how it is possible to secure the best value from such a service under the circumstances. The fact that at least one CCG managed to achieve reasonably good coverage indicates the possibility for learning from others.

## Data Availability

Individual patient-level data and data supplied under specific data sharing agreements cannot be made available by the study team. Sources for data that are already publicly available are supplied either in the text or the references. Aggregate survey data collected by the study team will be presented when findings from the relevant workstreams to which they correspond have been published. Other data produced in the study are available upon reasonable request to the authors.

## Conclusion

This study provides an evaluation of the national implementation of remote home monitoring of pulse oximetry for people diagnosed with COVID-19 across the English NHS. Although we detected no impact on outcomes, there are potential explanations for this finding that are unrelated to the effectiveness of the programme. Taking due account of populations that may respond less well to oximetry, there is no evidence that future implementation of similar programmes would be unsafe. However, the challenges of providing sufficient data so that effectiveness can be adequately measured need to be overcome.

## Acknowledgements

The authors would like to thank the following: our NIHR BRACE and NIHR RSET public patient involvement members; Russell Mannion for peer-reviewing our study protocol; Public Health England and the Institute of Global Health Innovation, NIHR Patient Safety Translational Research centre, Imperial College London, NHS England and Kent, Surrey and Sussex Academic Health Sciences Network for providing data; Dr Cono Ariti (University of Cardiff) for statistical advice; and Eilis Keeble (Nuffield Trust) for extracting Hospital Episode Statistics.

We thank the NHS Digital CO@h Evaluation Workstream Group chaired by Professor Jonathan Benger for facilitating and supporting the evaluation, and to the other two evaluation teams for their collaboration throughout this evaluation: i) Institute of Global Health Innovation, NIHR Patient Safety Translational Research centre, Imperial College London and ii) the Improvement Analytics Unit (Partnership between the Health Foundation and NHS England and NHS Improvement).

We would also like to thank our Clinical Advisory Group for providing insights and feedback throughout the project (Dr Karen Kirkham (whose previous role was the Integrated Care System Clinical Lead, NHSE/I Senior Medical Advisor Primary Care Transformation, Senior Medical Advisor to the Primary Care Provider Transformation team), Dr Matt Inada-Kim (Clinical Lead Deterioration & National Specialist Advisor Sepsis, National Clinical Lead - Deterioration & Specialist Advisor Deterioration, NHS England & Improvement) and Dr Allison Streetly (Senior Public Health Advisor, Deputy National Lead, Healthcare Public Health, Medical Directorate NHS England).

## Contributors’ statement

CSJ, TG, SM, SMT and EM contributed to study design and methodology. CSJ and TG participated in data curation, analysis and validation. CSJ and TG have verified the underlying data. NJF led the overall mixed methods evaluation of which this study relates to one workstream. All authors provided input from their own workstreams either with raw data or to help interpret findings. CSJ led on the preparation and writing of the manuscript. All authors reviewed and provided feedback on the manuscript and approved the final version.

## Data sharing statement

Individual patient-level data and data supplied under specific data sharing agreements cannot be made available by the study team. Sources for data that are already publicly available are supplied either in the text or the references. Aggregate survey data collected by the study team will be presented when findings from the relevant workstreams to which they correspond have been published.

