## Supplementary file for "The impact of remote home monitoring of people with COVID-19 using pulse oximetry: a national population and observational study"

### Supplementary material

#### Contents

1. Data sources
2. Judging completeness of data within CCGs
3. Estimating exposure components of the regression models
4. Sensitivity analysis

### 1. Data sources

Table 1 provides details of the different data sources used in the study.

**Table 1: Sources of data and information used in the study**

| Data | Source | Details |
|---|---|---|
| Mortality within 60 days of first laboratory-confirmed case or with confirmed COVID-19 present on death certificate | Public Health England (Now UK Health Security Agency) | By age band, CCG, week |
| New cases of laboratory-confirmed Covid-19 | Public Health England | By age band, CCG, week |
| People onboarded to CO@h | NHS Digital: bespoke data collection from the programme aggregated by Imperial College London | By age band, CCG and fortnight, rounded to the nearest five patients or labelled as between one and seven. |
| Hospital admissions for Covid-19 or suspected Covid-19 | Hospital Episode Statistics (HES) | Individual patient-level data aggregated by age band, fortnight and CCG of responsibility |
| In-hospital mortality | HES | Individual patient-level data |
| Lengths of hospital stay | HES | Individual patient-level data |
| Patient characteristics on admission | HES | Individual patient-level data |
| The proportion of acute beds occupied patients with Covid-19 | NHS England and NHS Improvement | By acute trust, daily |
| The presence of a post-discharge Covid virtual ward | Kent, Surrey and Sussex Academic Health Sciences Network | By acute trust |

### 2. Judging completeness of data within CCGs

We combined two sources of information to judge completeness of data:

(i) The management information collected by NHS Digital from each site;
(ii) Onboarding data received by the programme; and
(iii) Replies to the costing survey administered by the study team and sent to 28 sites.

The management information provided assessments as to whether the data reported by each site was complete up to mid-April 2021, the onboarding data covered the period from October 2020 to the end of April 2021 and the survey asked for numbers of individuals onboarded from the date the service started up to the end of April 2021.

For the 28 sites included in the survey, we compared the total numbers of onboarded individuals in the data we received from the programme (the programme data) to the numbers reported in the survey.

For most sites the numbers were broadly similar. However, among the CCGs reported as complete in the management information we excluded three CCGs where the numbers onboarded in the programme data were below 60% of those in the survey. We also included three CCGs where the data was not reported as complete but the numbers recorded as onboarded within the programme data were approximately the same as, or exceeded the numbers in the survey.

#### 3. Estimating exposure components of the regression models

For our modelling of mortality and hospital admission we required estimates of exposure to COVID-19 so that we could then relate rates of outcome to levels of coverage and other variables. For example, for mortality, the basic regression model used is:

$$Log(number\ of\ deaths(t)) = Log(Exposure(t)) + \beta_0 + \beta_1(Coverage(t)) + \beta_2(Age\ band) + \beta_3(Month)$$

Where the $\beta_i$'s are regression coefficients and *t* denotes the fortnight.

A simple approach would be to estimate exposure as the number of new cases in the same period as the deaths occurred, but, given many of those dying would have been identified as new cases some weeks before, this is unrealistic and would overestimate the exposure while cases are rising and underestimate it when cases are falling. A better approach would be to recognise the median time between diagnosis and death as about two weeks, and so use the number of new cases in the previous fortnight. In our study we went a further step and implemented an approach that applied weights to the case data from more than one previous time period. These weights reflect the relative contributions of each time period, sum to one, and can be estimated by linear regression, assuming the relationship remains constant over the period of the analysis (see Figure 1).

**Figure 1: The application of weights to the current and previous time periods (fortnights) to create the exposure associated with outcomes. The sum of weights: $w_0 + w_1 + w_2 = 1$**

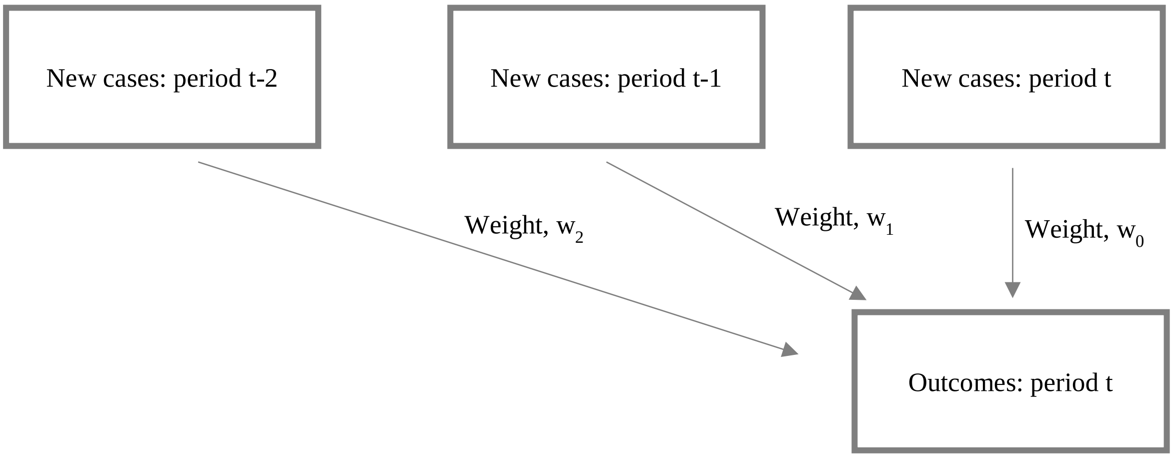


Assuming onboarding into the CO@h programme occurs soon after diagnosis, the lags and corresponding weights used for the onboarding data remain the same. The weighted onboarding numbers divided by the weighted new cases then becomes the coverage value that is used in the final regression model shown above.

The weights that we used are shown in Table 2. If we included lags of more than two fortnights, the estimated weights for those periods became very small and lacked statistical significance, so we carried out our final estimates by only going back as far as two previous fortnights. Different weightings were selected for the sensitivity analysis to see how they affected results.

**Table 2: Weights applied to lagged numbers of new cases for each outcome. ($w_0$ is applied to new cases in the same period as the outcome is measured, $w_1$ is applied to new cases in the previous fortnight and $w_2$ to the fortnight before that.)**

| | | Weight | | |
|---|---|---|---|---|
| **Outcome** | **Age band** | **$w_0$** | **$w_1$** | **$w_2$** |
| Mortality | 65 to 79 | 23.1% | 60.2% | 16.6% |
| | 80+ | 27.5% | 67.4% | 5.0% |
| Hospital admission | 65 to 79 | 61.1% | 37.8% | 1.1% |
| | 80+ | 81.8% | 14.5% | 3.7% |

.

### 4. Sensitivity analysis

For sensitivity analysis we tested different scenarios for weighting lagged variables to create different values for exposure in our regression models. We also investigated outcomes if we excluded hospital admissions for suspected COVID-19, focussing exclusively on confirmed diagnoses. For the weighting scenarios we chose the same weighting for both age bands and varied them across a range of feasible values. For the in-hospital outcomes the weightings are applied to the coverage and correspond to those for admissions.

Under each scenario, the impacts of a 10% increase in coverage on each outcome are shown in Tables 3 to 5. None of the effects are statistically significant at the 5% level (two-sided), although the impact on the risk of hospital admission without any lags ($w_0$ = 100%, $w_1$ = 0%, $w_2$ = 0%), or with a lag of just one fortnight ($w_0$ = 0%, $w_1$ = 100%, $w_2$ = 0%) are borderline significant for a positive relationship (p=0.06 in both scenarios).

**Table 3: The impact of coverage on the risk of mortality under different modelling assumptions**

| **Scenario** | | **Relative risk of death associated with a 10% increase in coverage *(95% confidence interval)*** | |
|---|---|---|---|
| Baseline | (see table 2) | 0.98 | *(0.96, 1.01)* |
| Weighting (applied to both age bands) | $w_0$ = 30%, $w_1$ = 50%, $w_2$ = 20% | 0.98 | *(0.95, 1.01)* |
| | $w_0$ = 10%, $w_1$ = 70%, $w_2$ = 20% | 0.98 | *(0.95, 1.01)* |
| | $w_0$ = 30%, $w_1$ = 70%, $w_2$ = 0% | 1.00 | *(0.97, 1.03)* |
| | $w_0$ = 0%, $w_1$ = 100%, $w_2$ = 0% | 1.00 | *(0.97, 1.02)* |

.

**Table 4: The impact of coverage on the occurrence of hospital admission under different modelling assumptions**

| **Scenario** | | **Relative risk of admission associated with a 10% increase in coverage *(95% confidence interval)*** | |
|---|---|---|---|
| Baseline | (see table 2) | 1.03 | *(0.99, 1.07)* |
| Weighting (applied to both age bands) | $w_0$ = 60%, $w_1$ = 40%, $w_2$ = 0% | 1.02 | *(0.98, 1.06)* |
| | $w_0$ = 100%, $w_1$ = 0%, $w_2$ = 0% | 1.03 | *(1.00, 1.07)* |
| | $w_0$ = 0%, $w_1$ = 100%, $w_2$ = 0% | 1.05 | *(1.00, 1.10)* |
| | $w_0$ = 50%, $w_1$ = 50%, $w_2$ = 0% | 1.02 | *(0.98, 1.06)* |
| | $w_0$ = 60%, $w_1$ = 30%, $w_2$ = 10% | 1.01 | *(0.97, 1.05)* |
| Exclude patients with suspected COVID-19 as primary diagnosis | | 1.01 | *(0.97, 1.04)* |

.

**Table 5: The impact of coverage on in-hospital mortality and length of stay under different modelling assumptions**

| **Scenario** | **Odds ratio associated with in-hospital mortality for every 10% increase in coverage** ***(95% confidence interval)*** | | **Relative change in length of stay for every 10% increase in coverage** ***(95% confidence interval)*** | |
|---|---|---|---|---|
| Baseline | 0.97 | *(0.92, 1.03)* | 1.8% | *(-1.2%, 4.9%)* |
| Weighting used to create coverage variable (applied to both age bands) | | | | |
| $w_0$ = 60%, $w_1$ = 40%, $w_2$ = 0% | 0.96 | *(0.91, 1.02)* | 1.7% | *(-1.4%, 4.9%)* |
| $w_0$ = 100%, $w_1$ = 0%, $w_2$ = 0% | 0.98 | *(0.93, 1.02)* | 1.2% | *(-1.3%, 3.7%)* |
| $w_0$ = 0%, $w_1$ = 100%, $w_2$ = 0% | 0.95 | *(0.90, 1.01)* | 0.9% | *(-1.7%, 3.6%)* |
| $w_0$ = 50%, $w_1$ = 50%, $w_2$ = 0% | 0.96 | *(0.90, 1.02)* | 1.7% | *(-1.4%, 5.0%)* |
| $w_0$ = 60%, $w_1$ = 30%, $w_2$ = 10% | 0.96 | *(0.91, 1.02)* | 2.1% | *(-1.1%, 5.4%)* |
| Exclude patients with suspected COVID-19 as primary diagnosis | 0.98 | *(0.92, 1.04)* | 0.2% | *(-2.8%, 3.3%)* |
